# Anticipating HIV Vaccines: A Cross-cultural Behavioral Economic Demand Analysis

**DOI:** 10.1101/2025.05.11.25327407

**Authors:** Promise Tewogbola, Oluwakamikun Faith Adekunle, Deborah Odaudu, Tolulope Oyewole

**Author notes:** Correspondence concerning this preprint should be addressed to Dr. Promise Tewogbola, Founder and CEO of PROMISE Labs Africa, a pioneering scientific nonprofit dedicated to cutting-edge translational behavioral research in Africa.

## Abstract

The development of effective vaccines remains crucial for controlling infectious diseases, with behavioral economics offering insights into vaccine acceptance patterns. This study aimed to understand demand for HIV vaccines currently in development, using behavioral economics, examining cross-cultural differences between Nigerian (*n* = 119) and United States (*n* = 63) participants. They responded to three behavioural economic tasks: a simulated COVID-19 Vaccine Purchase Task (CVPT), HIV Vaccine Purchase Task (HVPT), and the Monetary Probability Discounting Task (MPDT). Demand curves were fitted using an exponentiated model with both purchase tasks demonstrating excellent fit (*R*^2^ = .98). Multiple regression analyses examined relationships between behavioral economic indices and demographic factors. Nigerian participants showed significantly higher price sensitivity for both COVID-19 (*β* = −1.10, *p <* .001) and HIV vaccines (*β* = 0.90, *p* = .001) compared to U.S. participants, while baseline demand intensity and risk tolerance were similar between countries. Monetary risk-taking behavior predicted HIV vaccine demand (*β* = −1.10, *p* = .02), and higher socioeconomic status was associated with lower price sensitivity (*β* = −1.09, *p* = .04). The study demonstrates the utility of behavioral economic frameworks in modeling vaccine demand across cultures, revealing cross-cultural differences in price sensitivity and the potential relationship between COVID-19 and future HIV vaccine acceptance. These findings may have important implications for developing targeted public health interventions and policy frameworks for vaccine distribution, particularly in lower-income populations, to ensure equitable access to future HIV vaccines.

## 1. Introduction

Emerging and re-emerging infectious diseases continue to pose significant threats to global health. In the past 35 years alone, over 30 new human-affecting pathogens have emerged, including HIV/AIDS^1^, with most linked to zoonotic origins shaped by complex socioeconomic, environmental, and ecological factors (Nii-Trebi, 2017). Among available public health strategies, vaccination remains the most effective and safest method for preventing the spread of infectious diseases. According to the World Health Organization (2019a), vaccines prevent an estimated 2 to 3 million deaths annually.

Beyond direct disease prevention, vaccines provide a range of indirect benefits, including reductions in medical and socioeconomic burdens and transmission control through herd immunity. These community-level effects are particularly valuable for individuals unable to be vaccinated - such as infants, the elderly, or the immunocompromised (Chevalier-Cottin et al., 2020; Newell et al., 2010). Additionally, veterinary vaccines contribute to human health by reducing antimicrobial use in livestock, thereby slowing the emergence of antibiotic-resistant pathogens (Lekshmi et al., 2017).

However, despite these extensive benefits, vaccine confidence has eroded in some populations, fueled by misinformation and amplified by organized anti-vaccination movements. The now-debunked link between autism and the MMR vaccine is a prime example of how misinformation can undermine public health (Chevalier-Cottin et al., 2020; Dubé et al., 2021). In recognition of these challenges, the World Health Organization (2019b) identified vaccine hesitancy, the delay or refusal to vaccinate despite availability, as one of the top ten threats to global health.

Understanding and addressing vaccine hesitancy requires more than conventional epidemiological approaches. Behavioral economics offers a powerful interdisciplinary framework for examining how individuals make health-related decisions, integrating psychologically realistic assumptions into economic models of behavior (Angner & Loewenstein, 2012; Thaler, 2018; Hursh & Roma, 2013). Within this framework, two constructs, demand and discounting, are particularly useful in modeling vaccine acceptance.

Demand refers to the relationship between the quantity of a health commodity consumed (e.g., a vaccine) and its associated cost. This relationship typically follows a nonlinear, downward-sloping demand curve (Allen, 1938; Jacobs & Bickel, 1999), as shown in Figure 1. Importantly, in behavioral economics, “cost” can extend beyond monetary terms to include time, effort, side effects, or perceived vaccine inefficacy. The willingness-to-pay (WTP) approach formalizes this by capturing how much individuals value a good across a range of perceived costs (Phelps, 2017).

**Figure 1:**
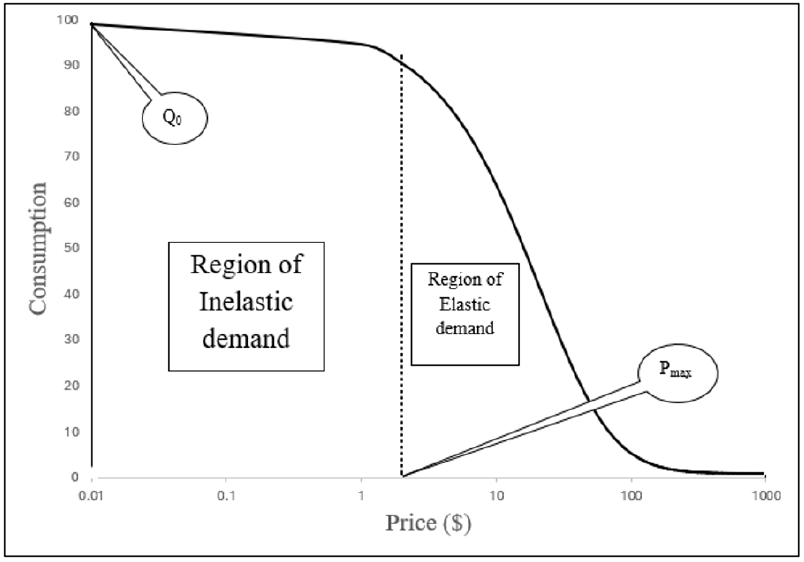
Schematic exponential demand curve plotted with price on logarithmic axes. *Note. Q*_0_ depicts the highest level of consumption when the commodity is free. Demand transitions from inelastic to elastic at *P*_max_, which is the price point corresponding to where the slope of the demand curve is −1

To quantify and analyse demand more precisely, behavioral economists often model demand curves using the exponentiated demand equation (Koffarnus et al., 2015):

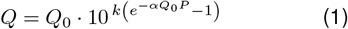

Here, *Q* represents the quantity of the commodity of interest consumed, and *Q*_0_ depicts the demand intensity, which is the highest level of demand when that commodity is free. The parameter *α* specifies the rate of sensitivity of consumption to increases in cost. *P* stands for price (broadly defined), and *k* is the scaling constant reflecting the range of consumption data in log units. Among these, *Q*_0_ and *α* are two of the most important indices used in comparing demand between individuals or across population groups. *Q*_0_, representing the highest level of demand at zero or extremely low costs, provides crucial insights into vaccine acceptance. A high *Q*_0_ implies that a respondent is very likely to accept a vaccine as long as the cost is at or approaches zero. This can indicate a strong baseline willingness to be vaccinated when the barriers are minimal. On the other hand, *α*, which reflects price sensitivity, offers information about how demand changes as cost increases. A more negative *α* suggests that the respondent will be relatively insensitive to higher costs in order to access a vaccine. In other words, these individuals might be willing to pay more or overcome greater barriers to obtain the vaccine. While demand analysis offers valuable insights into decision-making by examining how consumption changes as a function of cost, it is notable for its flexibility in how “cost” can be defined.

However, vaccine decisions, like many health behaviors, involve considerations of delayed and uncertain outcomes. The behavioral economic concept of *discounting* addresses these dimensions. *Probability discounting*, specifically, examines how the subjective value of an outcome diminishes as the uncertainty or risk associated with obtaining it increases (Madden & Bickel, 2010; Rachlin, 2004). Understanding individual differences in probability discounting (e.g., risk aversion vs. risk seeking) can provide critical insights into vaccine acceptance, where perceived efficacy and potential side effects introduce elements of uncertainty.

Figure 2 shows two nonlinear curves representing discounting in individuals whose behavior would be described as “risk avoidant” or “risk seeking,” depending upon the relative degree of discounting as a function of odds against receiving the commodity.

**Figure 2:**
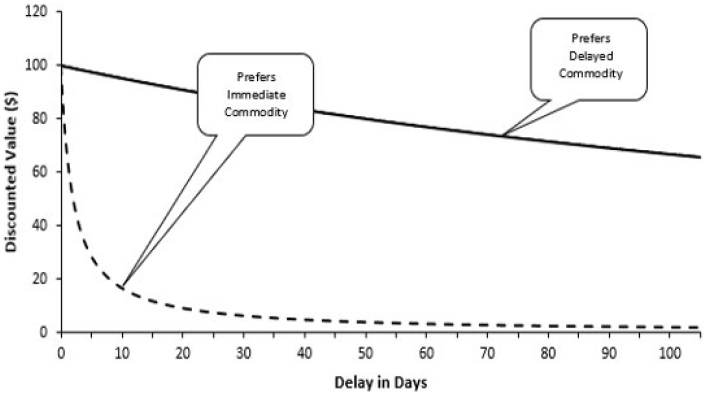
Prototypical discounting curves demonstrating the decay in value of $100 as a function of odds against its receipt. *Note*. Graph represents how two individuals discount the value of a monetary reward accessible after a certain delay. Note how the smooth, continuous curve shows a gentle decline in the value of $100 with increasing odds against receiving it, while the decline is more rapid in the dashed curve. This implies that the individual with the dashed curve is more intolerant of uncertainties in accessing the monetary reward compared to the individual with the smooth continuous curve.

Probability discounting can be quantified using a two-parameter hyperboloid function (Rachlin, 2006):

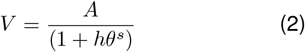

Here, *V* represents the subjective value of the risky or uncertain commodity, while *A* is the amount of the risk commodity. The term *θ* denotes the odds against accessing the uncertain commodity. The parameter *h* reflects the discounting rate, which is crucial in comparing discounting between individuals or groups. Finally, *s* represents the sensitivity to uncertainty.

In discounting experiments, the discounting parameter *h* is typically treated as a dependent variable. High values of *h* indicate risk aversion, while low values suggest risk-seeking tendencies. The parameter *s* captures the sensitivity to uncertainty in the context of probability discounting. Higher values of *s* indicate a greater impact of uncertainty on the subjective value of the commodity.

In addition to these theoretical models, Myerson et al. (2001) introduced an atheoretical approach utilizing Area Under the Curve (AUC) to measure behavioral economic indices. This method provides a model-free alternative for quantifying both demand and discounting. The process involves normalizing price (*P*) or odds (*θ*), and the value to which consumption (*Q*) or amount (*A*) is set, so they span from 0 to 1. AUC is then determined by summing the areas of the trapezoids formed between each interval, calculated using the following equation:

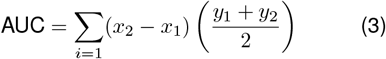

where *x*_1_ represents the normalized smaller interval value, *x*_2_ the normalized larger interval value, and *y*_1_ and *y*_2_ are the corresponding normalized consumption or amount points. The aggregate of these areas provides a singular AUC value per individual. AUCs range from 0 to 1, with larger AUC values signifying less discounting or higher demand, while smaller AUC values connote more pronounced discounting or lower demand.

This consideration is particularly urgent in the context of HIV vaccine development. Despite on-going research, no effective HIV vaccine has been rolled out to the public. However, recent advances in mRNA technology, spurred by COVID-19 vaccine development, have accelerated progress (Harris, 2022; Rogers, 2022). Furthermore, anticipating demand for vaccines still in development, such as for HIV, is crucial for informing public health preparedness and ensuring effective access and acceptance upon their availability (Levine et al., 2008). This study leverages Hypothetical Purchase Tasks to conduct a pioneering behavioral economic analysis of HIV vaccine acceptance in Nigeria and compares it to the United States. Understanding potential demand in diverse populations before market release can significantly enhance targeted public health initiatives and policy development.

With these considerations in mind, the present study has three main goals: (1) evaluate the utility of behavioral economic models in describing demand for HIV and COVID-19 vaccines. We hypothesize that participant responses on both the HIV Vaccine Hypothetical Purchase Task (HVPT) and COVID-19 Vaccine Hypothetical Purchase Task (CVPT) will closely resemble the prototypical downward-sloping curve observed in behavioral economic studies and be described by the exponentiated demand equation (Koffarnus et al., 2015); (2) investigate cross-cultural differences between Nigeria and the United States in three behavioral economic measures: the CVPT, HVPT, and Monetary Probability Discounting Task (MPDT). This analysis will provide insights into how cultural factors may influence vaccine acceptance and risk behavior across these two diverse populations; and (3) explore the potential to predict demand for the HIV vaccines currently in development. Specifically, the study aims to examine how demand for HIV vaccines may be predicted by demand for COVID-19 vaccines, which could suggest that vaccination behavior follows habitual patterns. Additionally, the relationship between risk behavior, as measured by the MPDT, and HIV vaccine demand, will be investigated acknowledging the complex interplay between risk tolerance in monetary domains and health-related decision-making. Finally, analysis will be conducted on how demographic factors such as age, gender, geographic location, and socioeconomic status may predict demand for HIV vaccines. With this analysis, the study aims to provide valuable insights into the determinants of vaccine demand across cultures, informing the development of targeted public health interventions and policy formulations for both COVID-19 and future HIV vaccines.

## 2. Methodology

### 2.1. Participants

#### 2.1.1 Procedure

The Institutional Review Board (IRB) at a State University in the midwestern region of the United States approved all study procedures. American participants were obtained through Amazon’s crowdsourcing platform, Mechanical Turk (MTurk), while Nigerian participants were sourced from social media websites. Participants chose to voluntarily engage in this study by clicking on a link which directed them to a Qualtrics-hosted web survey. The first page of the survey presented an informed consent form, and participants continued if they agreed to participate. American participants were compensated with $1 for their participation, consistent with standard MTurk compensation rates for tasks of similar length and complexity. Due to logistical constraints and the nature of recruitment, Nigerian participants were not offered any compensation. This differential treatment is acknowledged as a significant limitation of the study and is discussed further in the limitations section.

### 2.2. Measures

#### Demographics

Participants were asked to provide demographic information including their gender, age, and socioeconomic status. For gender, participants could select from options of “female,” “male,” or “other.” Age was collected as a numeric value. Socioeconomic status was self-reported with participants selecting one of the following categories: “lower,” “working,” “middle,” “upper middle,” and “upper” class. Additionally, a “Country” variable was created based on the recruitment source: participants recruited from the United States were coded as “USA,” while those recruited from Nigeria were coded as “NGA.”

#### COVID-19 Vaccine Purchase Task (CVPT)

This is a simulated purchase task adapted from Hursh et al. (2020) and used to assess the likelihood of accepting a COVID-19 vaccine at different risks of hospitalization due to COVID-19. The following instructions were provided:

Please read and consider the following. Vaccines provide immunity to a disease by developing antibodies to fight it – this means that while you may still get COVID-19 if vaccinated, the vaccine can reduce the chance of hospitalization at different levels of vaccine efficacy. You can get the vaccine through your doctor, at no cost to you. Imagine you have not been vaccinated yet and a COVID-19 vaccine has been approved by the XDA only after it has undergone a standard and rigorous vaccine evaluation. This evaluation included all 3 phases of human clinical trials to determine the vaccine’s safety and effectiveness. You can get the vaccine through your doctor, at no cost to you. Assumptions: The vaccine is easily administered; The vaccine is available to you without cost (free); You have the same income/savings as you do now; You have no access to any other vaccines available for COVID; The vaccine must be administered at the time of receiving it (you can’t save it to use at a later date); This vaccine must only be used for you (you cannot use this vaccine for friends or family members); This vaccine is approved by XDA. Remember that there are no “right” or “wrong” answers. Please respond honestly, as if you were actually in this scenario. Given the above scenario, please slide the bar to indicate the likelihood of you getting the COVID-19 vaccine if the pharmaceutical company claims it reduced chances of hospitalization by each of the following percentages (X%).

The value of XDA indicates the national regulatory body for food and drugs – the Food and Drug Administration (FDA) for US participants or the National Agency for Food and Drug Administration and Control (NAFDAC) for Nigerian participants. Participants were then asked to use a slider on a visual analogue scale (VAS) to indicate their willingness to accept a vaccine at various risks of hospitalization due to COVID-19. Vaccine efficacy was described as the percentage reduction in risk of hospitalization on account of COVID-19 vaccination. Vaccine efficacy ranged from 100% (totally effective) to 0% (totally ineffective) in decrements of 10 percentage points.

#### HIV Vaccine Purchase Task (HVPT)

This task was identical in structure and administration to the CVPT, with the only difference being that it assessed the likelihood of accepting an HIV vaccine at different risks of HIV infection. The same range of vaccine efficacy (100% to 0%) and instructions were used, with the relevant adjustments for HIV in place of COVID-19.

#### Monetary Probability Discounting Task (MPDT)

This measure was used to assess risk behavior through the probability discounting of hypothetical monetary rewards. The following instructions were provided:

Please read and consider the following. In the following decision-making task, you will be asked how much (in hypothetical M) you are willing to collect for certain to forgo an X% chance of obtaining Y. Remember that there are no “right” or “wrong” answers. Please respond honestly, as if you were actually in this scenario. In the scenarios that follow, please drag the slider to the value that corresponds with your choice.

The value of M represents the local currency: dollars ($) for US participants and naira 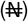 for Nigerian participants; X represents the varying likelihoods of obtaining a risky large monetary reward and included probabilities of 90%, 75%, 50%, 25%, and 10%; Y represents the risky large monetary reward, which was $1000 for US participants or 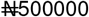 for Nigerian participants.

### 2.3. Statistical Analysis

A two-stage approach was employed for the analysis of demand. In the first stage, subject-specific demand indices (*Q*_0_ and *α* values) were estimated through nonlinear regression by fitting the exponentiated demand function (Koffarnus et al., 2015) to each participant’s data. Individual *α* values were log-transformed due to their non-normal distribution. For the discounting analysis, discounting rates for each participant were initially estimated using both the AUC method and Rachlin’s (2006) hyperbolic discounting model with parameters *h* and *s*. In the second stage, multiple linear regression analyses were conducted to examine the variability in the behavioral economic indices (*Q*_0_, *α*, demand AUC, *h, s*, and discounting AUCs). Due to the non-normal distribution of all indices, a logarithmic transformation was applied before analyzing variations in these values. To account for multiple comparisons and reduce the risk of Type I errors, a Bonferroni correction was applied. The significance level was adjusted by dividing the conventional p-value of 0.05 by the number of tests (16), resulting in a corrected significance threshold of approximately *p <* 0.003 (0.05/16). All statistical analyses were performed using R version 4.1.0 (R Core Team, 2021).

## 3. Results

### 3.1. Participant Demographics

A total of 182 participants were included in the study. The average age was 33.33 years (*SD* = 10.05). In terms of gender, 15.93% identified as female (*n* = 29), 32.97% as male (*n* = 60), and 50.99% (*n* = 93) either preferred not to answer or did not respond. Most participants were based in Nigeria (65.38%, *n* = 119), with the remaining 34.62% (*n* = 63) residing in the United States. Self-reported socioeconomic status varied, with 1.10% identifying as lower class (*n* = 2), 19.78% as working class (*n* = 36), 24.18% as middle class (*n* = 44), 3.85% as upper-middle class (*n* = 7), and 0.55% as upper class (*n* = 1). However, 50.55% (*n* = 92) did not provide information on socioeconomic status. Full demographic details are presented in Table 1.

**Table 1:** Demographic Information of Study Participants.

| Variable | Count / Mean | % / SD |
| --- | --- | --- |
| **Age** | 33.33 | 10.05 |
| **Gender** |  |  |
| Female | 29 | 15.93% |
| Male | 60 | 32.97% |
| Prefer Not to Answer / Did Not Answer | 93 | 50.99% |
| **Country** |  |  |
| Nigeria | 119 | 65.38% |
| USA | 63 | 34.62% |
| **Socioeconomic Status** |  |  |
| Lower | 2 | 1.10% |
| Working | 36 | 19.78% |
| Middle | 44 | 24.18% |
| Upper | 1 | 0.55% |
| Upper Middle | 7 | 3.85% |
| Did Not Answer | 92 | 50.55% |

### 3.2. Evaluation of Behavioral Economic Models

Responses elicited with the hypothetical purchase tasks produced demand curves that demonstrated excellent fit to the empirical data regardless of vaccine type. Group-level analyses revealed strong fits for both vaccine purchase tasks (COVID-19: *R*^2^ = .98; HIV: *R*^2^ = .98). Individual-level analyses showed similarly robust fits, with median *R*^2^ values of .85 (IQR: .55–.91) for COVID-19 vaccines, and .87 (IQR: .31–.92) for HIV vaccines. Aggregate demand curves across all participants are shown in Figure 3. Figure 4 and 5 presents box plots stratifying the demand indices by country and vaccine type.

**Figure 3:**
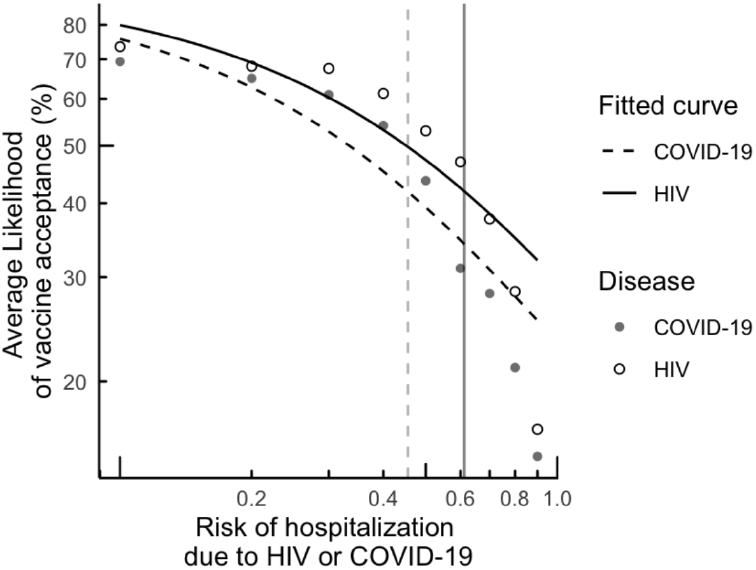
Aggregate Vaccine Demand Curves for HIV and COVID-19 as a Function of Hospitalization Risk *Note*. This figure shows aggregated vaccine demand curves for HIV and COVID-19 plotted on logarithmic scales, where the price is represented by the perceived risk of future hospitalization due to either disease. The continuous line represents demand for HIV vaccines, while the dashed line represents demand for COVID-19 vaccines. The plotted points reflect average likelihood of vaccine acceptance across levels of perceived risk. *P*_*max*_ values (vertical lines) mark the transition from inelastic to elastic demand, calculated using the Lambert W function based on fitted demand curve parameters *Q*_0_ and *α* (Gilroy et al., 2019). Aggregate demand for HIV vaccines was consistently higher than for COVID-19 vaccines across the risk spectrum.

**Figure 4.**
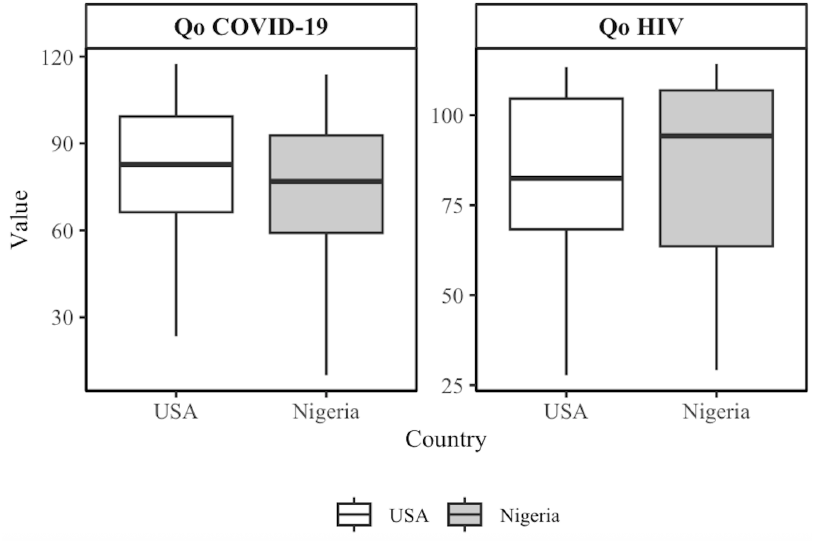
Boxplots depicting *Q*_0_ values for COVID-19 and HIV vaccines segregated by country. Horizontal black lines represent the median. Whiskers are 1.5 x IQR. Higher values indicate higher demand intensity. For COVID-19 vaccines, Nigerian participants exhibited a lower median *Q*_0_ than U.S. participants. In contrast, for HIV vaccines, the median *Q*_0_ was higher among Nigerian participants than among their U.S. counterparts.

**Figure 5.**
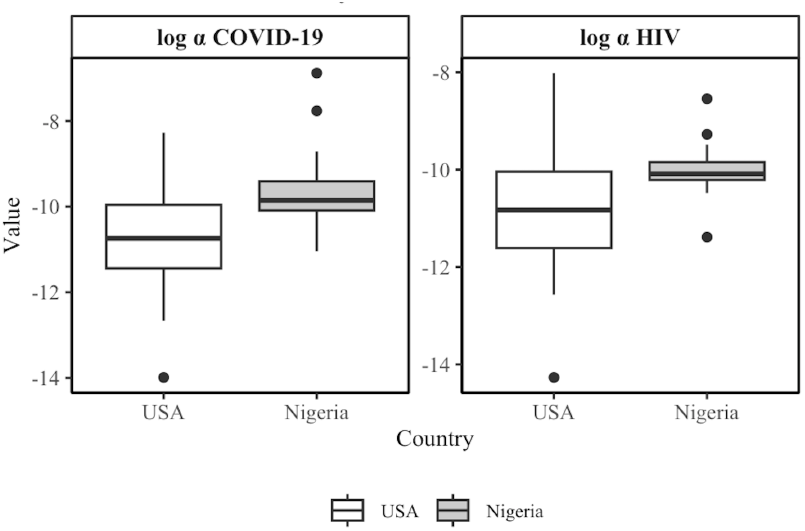
Boxplots depicting *α* values for COVID-19 and HIV vaccines segregated by country. Horizontal black lines represent the median. Whiskers are 1.5 x IQR. Higher values indicate higher price sensitivity.

The monetary probability discounting data also demonstrated the characteristic hyperbolic shape, with individual *R*^2^ values being notably high (median *R*^2^ = .96, IQR = .74 to .99). Figure 6 shows aggregate probability discounting curves segregated by country.

**Figure 6.**
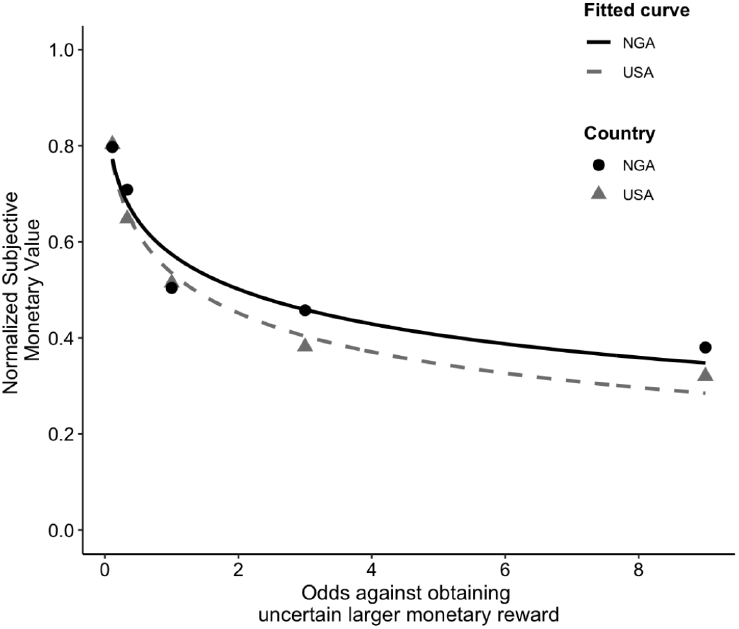
Aggregate Probability Discounting Curves for Nigerian and U.S. Respondents. *Note*. This figure depicts the devaluation of a monetary reward as a function of the odds against receiving it, based on aggregate probability discounting curves. The solid line represents aggregate data from Nigerian respondents (NGA), and the dashed line represents aggregate data from U.S. participants (USA). Data points reflect normalized subjective monetary reward values at each level of odds against its receipt. To enable cross-cultural comparison despite differences in currency denominations and exchange rates, monetary values were normalized prior to modeling. The fitted curves suggest that Nigerian participants, on average, displayed greater tolerance for probabilistic outcomes (i.e., less steep discounting) compared to their U.S. counterparts.

### 3.3. Cross-Cultural Differences

Linear regression analyses were conducted to examine cross-cultural differences between Nigeria and the United States in behavioral economic measures (Table 2). Nigerian participants demonstrated higher price sensitivity for both COVID-19 (*β* = 1.10, SE = 0.23, *p <* .001, *R*^2^ = .25) and HIV vaccines (*β* = 0.90, SE = 0.26, *p* = .001, *R*^2^ = .19). These differences remained significant after applying the Bonferroni correction, indicating that Nigerian participants’ demand for vaccines decreased more rapidly as price increased compared to U.S. participants. That said, no significant differences were found in demand intensity (*Q*_0_) or AUC for either vaccine task, suggesting similar baseline demand intensities between countries. Additionally, risk behavior, as assessed by monetary probability discounting, did not significantly differ between Nigerian and U.S. participants (*β* = −0.17, SE = 0.13, *p* = .186, *R*^2^ = .02).

**Table 2:**
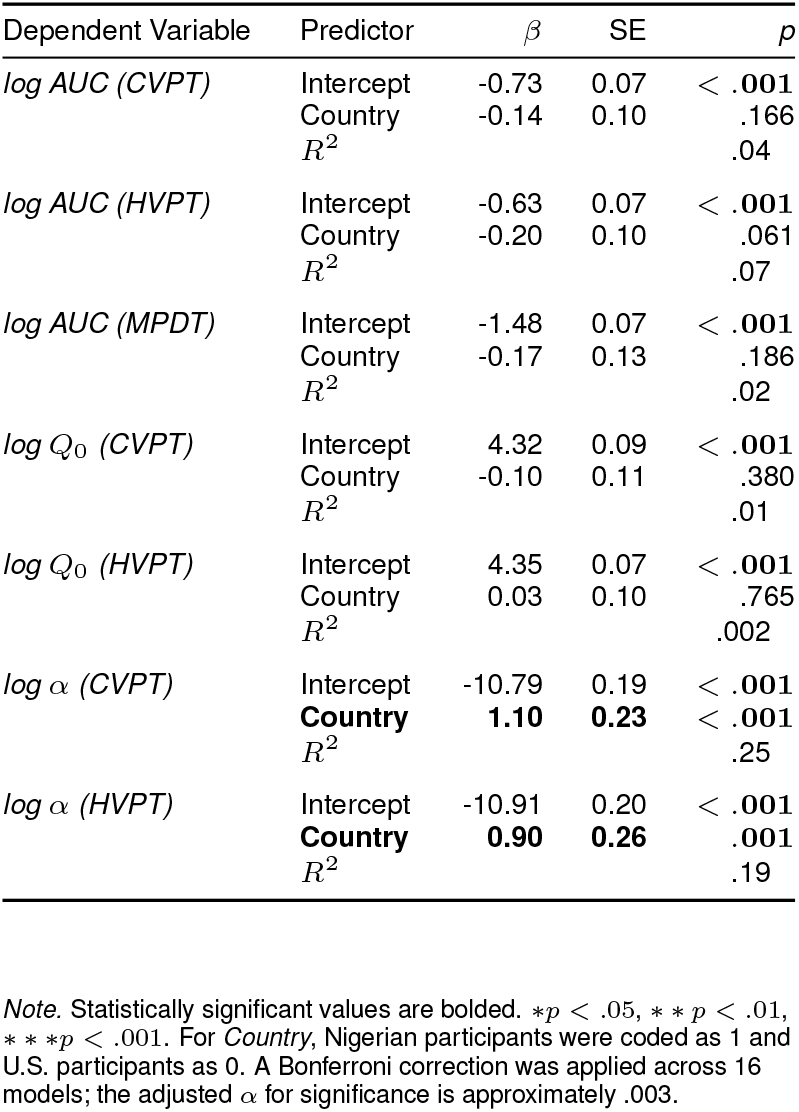
Linear Regression Analysis of BE Indices as a Function of Country.

### 3.4. Predicting HIV Vaccine Demand

Multiple linear regression analyses revealed several predictors of HIV vaccine demand parameters (Tables 3–5). COVID-19 vaccine demand indices strongly predicted corresponding HIV vaccine demand parameters. Specifically, COVID-19 vaccine demand intensity (*Q*_0_) significantly predicted HIV vaccine demand intensity (*β* = 0.99, SE = 0.17, *p <* .001, *R*^2^ = .676), and COVID-19 vaccine price sensitivity also significantly predicted price sensitivity of HIV vaccine (*β* = 1.01, SE = 0.15, *p <* .001, *R*^2^ = .823). Both relationships remained significant after the Bonferroni correction.

**Table 3:** Linear Regression Analysis of HIV Vaccine Demand AUCs as a Function of COVID-19 Vaccine Demand, Uncertain Monetary Rewards, and Demographic Factors.

| Dependent Variable | Predictor | $\beta$ | SE | $p$ |
| --- | --- | --- | --- | --- |
| *log AUC (HVPT)* | Intercept | -2.50 | 0.49 | < .001 |
|  | *log AUC (CVPT)* | -0.24 | 0.24 | .336 |
|  | *log Q<sub>0</sub> (CVPT)* | 0.09 | 0.08 | .277 |
|  | *log $\alpha$ (CVPT)* | **-0.13** | **0.04** | **.010** |
| | $R^2$ | | | .540 |
| *log AUC (HVPT)* | Intercept | -0.31 | 0.15 | .051 |
|  | ***log AUC (MPDT)*** | **0.19** | **0.08** | **.027** |
|  | *log h* | -0.03 | 0.03 | .320 |
|  | *log s* | 0.07 | 0.04 | .065 |
| | $R^2$ | | | .386 |
| *log AUC (HVPT)* | Intercept | -1.05 | 0.31 | .002 |
|  | *log Age* | 0.09 | 0.09 | .338 |
|  | *SES (lower)* | -0.04 | 0.06 | .439 |
|  | *SES (upper)* | 0.11 | 0.08 | .162 |
|  | *Gender* | 0.10 | 0.05 | .057 |
| | $R^2$ | | | .151 |

*Note.* Statistically significant values are bolded. \* $p < .05$ , \*\* $p < .01$ , \*\*\* $p < .001$ . For *Gender*, male participants were coded as 1 and all others as 0. For *SES (lower)*, participants identifying as Lower or Working class were coded as 1. For *SES (upper)*, participants identifying as Upper Middle or Upper class were coded as 1. The reference group was *SES (middle)*, coded as 1 for participants identifying as Middle class. A Bonferroni correction was applied for 16 models; adjusted $\alpha = .003$ .

**Table 4:**
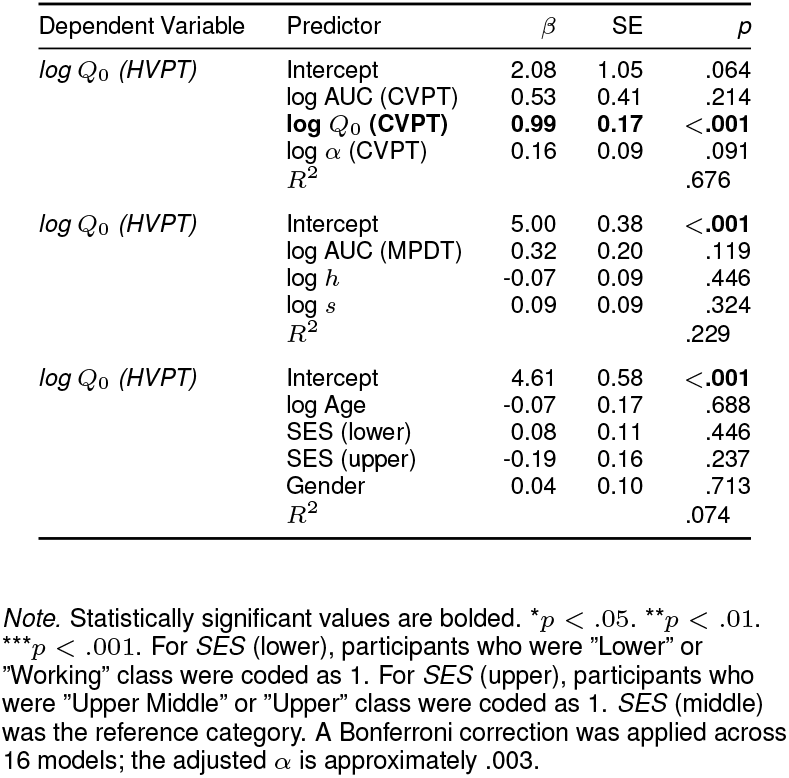
Linear Regression Analysis of HIV Vaccine Demand Intensity (Q_0_) as a Function of COVID-19 Vaccine Demand, Uncertain Monetary Rewards, and Demographic Factors.

**Table 5:**
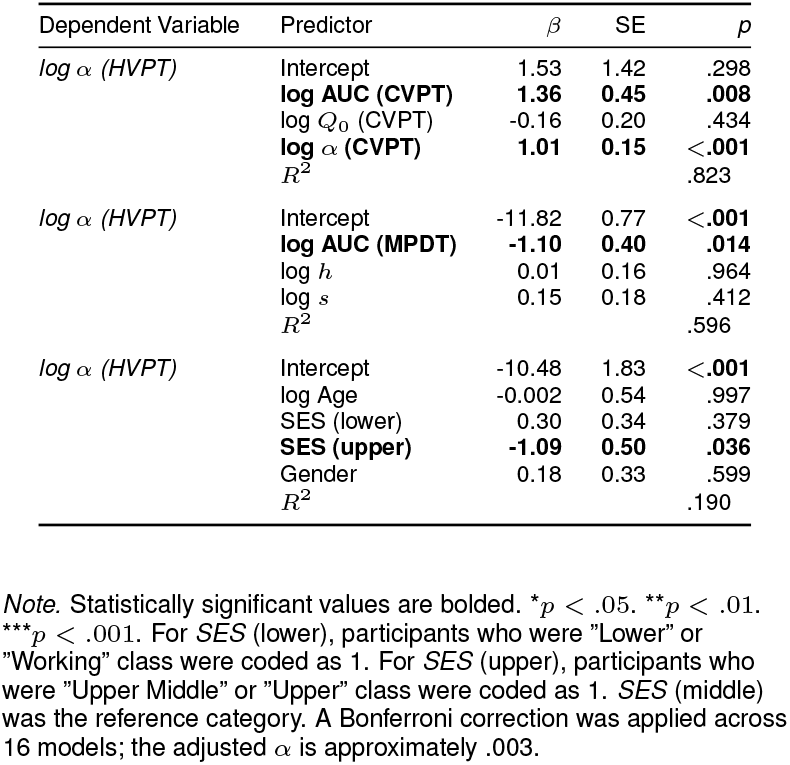
Linear Regression Analysis of HIV Vaccine Price Sensitivity (α) as a Function of COVID-19 Vaccine Demand, Uncertain Monetary Rewards, and Demographic Factors.

| Dependent Variable | Predictor | $\beta$ | SE | $p$ |
| --- | --- | --- | --- | --- |
| $\log \alpha$ (HVPT) | Intercept | 1.53 | 1.42 | .298 |
|  | **$\log$ AUC (CVPT)** | **1.36** | **0.45** | **.008** |
| | $\log Q_0$ (CVPT) | -0.16 | 0.20 | .434 |
|  | **$\log \alpha$ (CVPT)** | **1.01** | **0.15** | **<.001** |
| | $R^2$ | | | .823 |
| $\log \alpha$ (HVPT) | Intercept | -11.82 | 0.77 | **<.001** |
|  | **$\log$ AUC (MPDT)** | **-1.10** | **0.40** | **.014** |
| | $\log h$ | 0.01 | 0.16 | .964 |
| | $\log s$ | 0.15 | 0.18 | .412 |
| | $R^2$ | | | .596 |
| $\log \alpha$ (HVPT) | Intercept | -10.48 | 1.83 | **<.001** |
| | $\log$ Age | -0.002 | 0.54 | .997 |
|  | SES (lower) | 0.30 | 0.34 | .379 |
|  | **SES (upper)** | **-1.09** | **0.50** | **.036** |
|  | Gender | 0.18 | 0.33 | .599 |
| | $R^2$ | | | .190 |

Note. Statistically significant values are bolded. \* $p < .05$ . \*\* $p < .01$ . \*\*\* $p < .001$ . For SES (lower), participants who were "Lower" or "Working" class were coded as 1. For SES (upper), participants who were "Upper Middle" or "Upper" class were coded as 1. SES (middle) was the reference category. A Bonferroni correction was applied across 16 models; the adjusted $\alpha$ is approximately .003.

Additionally, monetary probability discounting significantly predicted price sensitivity of HIV vaccines (*β* = −1.10, SE = 0.40, *p* = .014), suggesting that individuals who valued probabilistic mone-tary rewards more showed less price sensitivity to HIV vaccine price increases. Socioeconomic status also emerged as a significant predictor, with individuals of higher socioeconomic status showing lower price sensitivity compared to those of middle socioeconomic status (*β* = −1.09, SE = 0.50, *p* = .036, *R*^2^ = .190). However, neither of these relationships survived the Bonferroni correction.

In predicting HIV vaccine demand AUC, COVID-19 vaccine price sensitivity was a significant predictor (*β* = −0.13, SE = 0.04, *p* = .010, *R*^2^ = .540), though this relationship did not remain significant after the Bonferroni correction. Monetary probability discounting AUC also significantly predicted the AUC for HIV vaccine demand (*β* = 0.19, SE = 0.08, *p* = .027, *R*^2^ = .386), indicating that a greater willingness to pay for probabilistic monetary rewards corresponded with higher HIV vaccine demand. However, this relationship also did not remain significant after the Bonferroni correction. Gender approached significance in predicting AUC for HIV vaccine demand (*β* = 0.10, SE = 0.05, *p* = .057, *R*^2^ = .151), suggesting a trend where male participants have a marginally higher demand for HIV vaccines compared to non-male participants.

## 4. Discussion

The primary objective of this study was to examine the dynamics governing vaccine demand, a crucial undertaking with significant implications for global public health. This research holds consid-erable importance, both conceptually and practically. Firstly, to our knowledge, this is among the pioneering studies employing a behavioral economic framework to specifically investigate the demand for a prospective HIV vaccine. Drawing on the success of prior vaccination campaigns against smallpox, polio, and most recently COVID-19, the promise of a preventive HIV vaccine is clear: long-term protection, reduced stigma, and cost-effective disease management compared to lifelong antiretroviral therapy (Duncan et al., 2012; Johnson & Neilands, 2007; Van Tam et al., 2011). The development of such a vaccine is no longer speculative, with mRNA-based HIV vaccine trials launched by the U.S. National Institute of Allergy and Infectious Diseases (Harris, 2022; Rogers, 2022; Stulpin, 2021), leveraging knowledge from the COVID-19 pandemic to accelerate HIV vaccine research. Thus, understanding the factors influencing the demand for a future HIV vaccine is both timely and necessary for its successful implementation and widespread adoption.

Secondly, this study represents a significant contribution by conducting behavioral economic analyses within a Nigerian population, a demographic often underrepresented in this field. While the principles of behavioral economics offer valuable insights into decision-making across various domains, the generalizability and robustness of these principles are enhanced by examining diverse populations, particularly those historically underserved by research. Although cross-cultural studies exist within behavioral economics (e.g., Chen, 2013; Doces & Wolaver, 2021; Domino, 1992; Henrich et al., 2010; Levinson & Peng, 2006; Wright & Phillips, 1980; Yates, 2002), the application to health behavior, particularly vaccine demand, remains limited. Our study addresses this critical gap by highlighting how cultural differences between Nigeria and the United States shape the demand for both existing (COVID-19) and prospective (HIV) vaccines.

Our findings demonstrate that demand for both COVID-19 and HIV vaccines can be accurately modeled using the exponentiated demand function, exhibiting the prototypical downward-sloping curve. The high model fits (*R*^*2*^ = 0.98 at the group level for both vaccines) highlight the strong explanatory power of the behavioral economic models in capturing vaccine demand, even when the cost is conceptualized as reduced vaccine efficacy rather than monetary expense. These results affirm the value of HPTs in capturing nuanced, quantitative insights into health decision-making and suggest that perceived efficacy functions similarly to price in shaping demand - an insight with important policy implications.

A central finding was that socioeconomic status (SES) significantly predicted price sensitivity for HIV vaccines, aligning with prior research indicating that individuals from lower socioeconomic backgrounds often exhibit heightened sensitivity to costs, whether financial or perceived (Jacob et al., 2022; Soofi et al., 2023; Wong et al., 2024). Given that socioeconomic status encompasses income, education, and occupation (Gonzalez et al., 2014), these results emphasize the importance of structural inequalities in shaping vaccine uptake (Caballo et al., 2021; Nfah-Abbenyi, 2024). The persistence of these socioeconomic differences in our study, even when ‘price’ was defined as vaccine inefficacy, highlights the broader influence of socioeconomic factors on health-related decision-making beyond mere affordability. This finding suggests that socioeconomic status affects health decisions through multiple pathways beyond direct financial constraints, potentially including differences in health literacy, risk perception, trust in healthcare systems, and opportunity costs associated with preventive healthcare.

Our cross-cultural analysis revealed that Nigerian participants exhibited significantly higher price sensitivity for both COVID-19 and HIV vaccines compared to their US counterparts, while no significant differences were observed in baseline demand intensity or risk tolerance as measured by the MPDT. This suggests that other factors besides differing risk profiles likely explain greater vaccine hesitancy in the Nigerian sample. This resonates with studies indicating that financial considerations may weigh more heavily in health decisions in Low- and-Middle-Income Countries (LMICs) (Nguyen et al., 2018), even when vaccines are free, as the “cost” is perceived in broader terms: potential ineffectiveness, time, access, or even opportunity costs. Beyond economic factors, differences in religiosity may also shape vaccine decision-making. Higher levels of religiosity in Nigeria (Pew Research Center, 2021; McKinnon, 2021) and the influence of religious leaders on health behavior (Heward-Mills et al., 2018) may contribute to vaccine hesitancy, especially when such leaders disseminate misinformation (The Lancet Global Health, 2023). However, as religiosity was not directly measured in this study, this interpretation warrants further investigation.

The absence of significant cross-cultural differences in risk tolerance may also suggest similarities in how risk is perceived. While this may appear inconsistent with prior research suggesting that individuals in developing economies are generally less risk-seeking (Vieider et al., 2012), it may reflect the homogenizing effects of globalization on psychological dispositions (Abdullahi, 2024), or a limitation of statistical power in this study.

The strong predictive relationship between COVID-19 vaccine demand indices and corresponding HIV vaccine demand parameters suggests a potential for behavioral consistency or ‘vaccination habituation’ (Fukui, 2022). Individuals who demonstrated a higher baseline willingness to accept a COVID-19 vaccine (higher *Q*_0_) were also more likely to exhibit a higher baseline willingness for an HIV vaccine. Similarly, those who were more sensitive to the perceived inefficacy of the COVID-19 vaccine were also more sensitive to the inefficacy of a hypothetical HIV vaccine. This pattern may indicate that prior experience with vaccination, even for a different disease, can normalize the process and reduce psychological barriers towards future vaccines (Mahameed et al., 2023). However, the generalizability of this effect is likely contingent on factors such as healthcare access (Lane et al., 2022), disease-specific risk perceptions (Hilverda & Vollmann, 2021), and trust in medical interventions (Lalumera, 2018). This insight has real-world value: vaccine attitudes may be shaped by prior behavior, implying that a successful rollout of one vaccine can improve acceptance of future ones, particularly among populations who have developed vaccine-seeking habits. Even when cost is conceptualized differently – such as uncertainty or efficacy – participants’ valuation of one vaccine translated to the other.

The observation that higher risk-seeking behavior (as indexed by AUC in probability discounting), is associated with increased demand for HIV vaccines carries significant implications for understanding vaccine adoption, although this relationship did not survive the stringent Bonferroni correction. This potential link suggests that individuals with a greater propensity for risk in monetary contexts might also be more inclined to accept the perceived uncertainties associated with vaccination as a preventive measure against HIV (Lepinteur et al., 2023). This could reflect a perception that the benefits of protection outweigh the potential risks, leading to more proactive engagement in health-promoting behaviors (Martín-Fernández & Ariza, 2016). However, this observation also raises concerns about a potential risk compensation or “moral hazard” (Moghtaderi & Dor, 2019), where vaccinated individuals increase risky behaviors under the false sense of security, particularly relevant in HIV prevention contexts.

Similarly, the finding that biological males exhibited a higher demand for HIV vaccines, while not statistically significant after correction, contrasts with observations during the COVID-19 pandemic where males often showed greater vaccine hesitancy (Otterbring & Festila, 2021; Steinhauer, 2021). This discrepancy warrants further investigation. Potential explanations could include differing perceptions of risk and susceptibility to HIV versus COVID-19, or gender-specific concerns related to vaccine side effects, particularly regarding fertility, pregnancy, and menstrual health, which were reported by some women following COVID-19 vaccination (Al-Husbain et al., 2023) and might influence their perception of future vaccines (Toshkov, 2023). Negative experiences with healthcare or difficulties accessing insurance, disproportionately affecting biological females in some contexts (Kakunami et al., 2008), could also contribute to this trend, although these factors were not directly assessed in our study.

Finally, age was not a significant predictor of vaccine demand in either country, contrary to assumptions that older individuals are more health-conscious or risk-averse (Oche et al., 2022). This finding suggests that other factors, such as socioeconomic status, access to healthcare, and health literacy, may exert a more dominant influence on vaccination decisions across diverse age groups. This has important implications for public health strategies, indicating that age alone may not be the most effective determinant for targeting vaccination campaigns. Instead, a more nuanced approach focusing on other demographic and psychosocial factors may be warranted. However, this null result may also reflect the limited power of our sample to detect more subtle age-related effects.

Our study possesses notable strengths. It is among the first to apply a behavioral economic framework to examine vaccine demand within the Nigerian population, offering novel insights into decision-making processes related to vaccine acceptance. The use of hypothetical purchase tasks (HPTs) allowed for a safe and controlled assessment of demand for a yet-to-be-experienced health commodity like an HIV vaccine in a Nigerian sample. Unlike dichotomous measures of willingness, HPTs provide a more nuanced understanding of demand by capturing responses across a range of hypothetical scenarios, enabling the isolation and control of factors influencing demand both between and within individuals (Strickland et al., 2022). This approach yields richer data for informing targeted public health interventions in Nigeria.

Nonetheless, limitations must be acknowledged. The use of non-probability sampling methods (MTurk for US participants and social media for Nigerian participants) introduces the potential for selection bias, which could affect the generalizability of our findings (Chandler & Shapiro, 2016). The social media recruitment in Nigeria presented challenges related to educational disparities, as the English-language survey may have disproportionately excluded individuals with lower levels of literacy, potentially leading to an overrepresentation of higher socioeconomic status individuals (Terasawa, 2024; UNESCO, 2002). The application of the Bonferroni correction, while necessary to control for Type I errors, increased the stringency of our significance threshold, potentially masking some true effects. Therefore, the observed findings, even those not reaching the adjusted significance level, should be considered as potentially informative for future research. The lack of compensation for Nigerian participants, due to logistical constraints, may have influenced participation rates and the representativeness of the Nigerian sample, potentially skewing it towards more altruistic or higher socioeconomic status individuals compared to the compensated US sample (Bierer et al., 2021; Kost & Correa, 2018). Finally, the reliance on MTurk for US participant recruitment carries inherent limitations associated with data quality concerns, including potential dishonesty and participant fatigue (Hauser et al., 2023; Moss et al., 2021). These methodological limitations should be considered when interpreting our findings.

## 5. Conclusion

This study provides new evidence that behavioral economic models are effective in predicting vaccine demand across cultural contexts, particularly for novel interventions like an HIV vaccine. Our results show that hypothetical demand is shaped by perceived efficacy, socioeconomic factors, and behavioral consistency across vaccine types. While vaccine price sensitivity varied across countries, with Nigerians more sensitive than Americans, the underlying behavioral relationships were stable, suggesting a general decision-making framework governs preventive health behavior. The identified limitations, including non-probability sampling and compensation disparities, underscore the need for methodological rigor in future cross-cultural research on health behaviors. Moving forward, it will be crucial to extend this work by exploring high-risk and underrepresented populations (e.g., MSMs, sex workers, migrant communities) and incorporating alternative cost metrics such as monetary price, side effects, or access barriers. Expanding cross-cultural behavioral economic research on health behaviors will be essential for designing effective, equitable vaccine strategies in the face of future pandemics and public health challenges.

## Data Availability

All data produced in the present study are available upon reasonable request to the authors

